# Machine Learning Algorithms to Fight the COVID-19 Pandemic

**DOI:** 10.1101/2022.12.31.22284091

**Authors:** Monalisha Pattnaik, Aryan Pattnaik, Alipsa Pattnaik

## Abstract

The ongoing novel COVID-19 global pandemic is one of the health emergencies in 21^st^ century after hundred years of Spanish flu that affected almost all the countries in the world. The objective of this study is to generate STM and LTM real-time out of sample forecasts of the future COVID-19 confirmed and death cases respectively for the top five mostly affected countries in the world namely USA, India, Brazil, Russia and UK. As of January 17, 2021, it has caused a pandemic outbreak with more than 94.5 million confirmed cases and more than 2 million reported deaths worldwide. Due to extreme robust behaviour in the univariate time series data, forecasting of both COVID-19 confirmed and death cases has become the exigent task for the government officials, healthcare workers, economists, corporate leaders, government, decision makers, public-policy makers, and scientific experts to allocate health resources. To solve this problem different hybrid approaches are applied which eliminate both linear and non-linear errors of the time series datasets and the predictions of for these countries will be practical to act as forewarning for all.

## 1. Introduction

The COVID-19 global pandemic is devastating and creating an uncertain situation in this 21^st^ century. Out of almost all countries in the globe USA, India, Brazil, Russia and UK are in the top to face this situation. In entire globe’s response to COVID-19 global pandemic, health and hospitals have played a vital role particularly for the countries like USA, India, Brazil, Russia and UK. It is essential to develop a forecasting model of COVID-19 for both confirmed and death cases of these five intensely affected countries.

On December 2019 for mystifying cause Wuhan city of China became the hub of an outbreak of pneumonia, latter it is named as COVID-19, which approached intense attention not only within China but globally (Gahan et al. 2021, Chakraborty et al., 2020, Guan et al., 2020, Ordonez et al., 2019 and Jung et al., 2020). This global pandemic is the most momentous global emergency which affects almost all the countries of the globe (Boccaletti et al., 2020).

On 28 Nov. 2020, it is reported worldwide (Nishiura et al., 2020) that outbreak has noted in a total of more than 6,171,5119 confirmed infections and more than 1,44,4235 confirmed deaths. On March 11 2020, World Health Organisation (WHO) declared COVID-19 as a “global pandemic” publicly, and then the United States declared it as a national emergency. Due to its widespread and contagious harm this unknown and unfinished COVID-19 has caused an obstacle to the health and safety of people all over the world. Thus, at this uncertain moment the continuous research of the COVID-19 pandemic and its prospect expansion has become a state-of-the-art research topic. Hence, it is necessary to develop the prediction model for forecasting of daily new confirmed and death cases for the intensely affected countries like USA, India, Brazil, Russia, and UK (Petropoulos et al., 2020). With the availability of limited and robust COVID-19 datasets, it is very difficult to predict this. From previous studies, it was evident that the timing and location of the outbreak facilitated the rapid transmission of the virus within a highly mobile population (Gahan et al. 2021, Roosa et al., 2020, Boldog et al., 2020 and Russel et al., 2020). In almost all affected countries, the governments imposed night curfew, a strict lockdown; patients who has the symptoms of COVID-19 are immediately quarantined or self isolation etc. For this situation of COVID-19 is putting a significant burden on the health care system and government of these countries. To fight this unfinished pandemic, various mathematical and statistical forecasting tools (Li et al., 2020 and Wu et al., 2020) and other resources are used. The authors (Fanelli et al., 2020, Kucharski et al., 2020) were developed to generate short-term and long-term forecasts of the confirmed cases. The single model prediction related to these datasets show wide range of fluctuations and inaccurate as the datasets of COVID-19 is robust and dynamic in nature. Hence, to predict and to generate STM and LTM out-of-sample forecasts hybrid modeling approach is developed. Previously traditional time series forecasting, like the ARIMA model is used frequently for forecasting univariate linear time series dataset (Box et al., 2015). To perform well to capture nonlinearity feature in time series data machine learning advanced models namely ANN, ARNN and WBF (Paul a al., 2017, Nury et al., 2017, Aminghafari et al., 2007, Beraouda et al., 2006 and Fay et al., 2007) are applied.

As COVID-19 data sets are usually both linear and nonlinear patterns so, using of single models like ARIMA or ARNN or SVM are not enough to predict. Therefore, the hybridization of both linear and nonlinear models can be well suited for precisely modelling of dynamic structures (Percival et al., 2020 and Chakraborty et al, 2020) to reduce the bias and variances of the prediction residuals of the component models. To solve a variety of time series issues shown in economic parameters, weather indicators, economic growth, GDP and wind speed etc. (Tumer et al., 2018, Kordanuli et al. 2017, Khashei et al., 2011, Milacic et al., 2017, Cadenas et al., 2010, Maleki et al. 2018 and Chakraborty et al., 2019, James et al., 2013 and Pattnaik et al. 2018) several hybrid approaches were developed in previous literature. To forecast dynamic time series accurately the hybrid ARIMA-ANN model (Zhang et al., 2003) has gained popularity due to its capacity. The selection of the number of hidden layers in the ANN architecture is the pitfall of this hybrid model that involves subjective judgment. This drawback may be identified by applying the ARNN model approach. It fits a feed-forward neural network model with only one hidden layer to a time series with lagged values of the series as inputs.

It provides less complexity, easy interpretability and better prediction as compared to WBF and ANN models in many situations (Hydmann, 2018). The practical intent of this real-time data-driven analysis is to provide health warriors, corporate leaders, economists, government/public-policy makers, and scientific experts with realistic estimates for the magnitude of the pandemic for policy-making for the dynamics of COVID-19 dissemination. To handle the dynamism and robustness of the characteristics of the COVID-19 time series data is the main motivation of this study. The most common solution of eliminating the dynamic properties of the time series datasets is to develop a hybrid model (Oliveira et al., 2014). Even from the researchers’ point of view, hybrid models are more efficient when all the data characteristics are not identified (Kuncheva, 2004). Through experimental evaluation, it is verified that the excellent performance of two proposed hybrid ARNNS and SVMAR models for both the confirmed and death cases respectively are shown for the global pandemics forecasting for intensely affected countries. Hence, these two models have easy interpretability, dynamic and robust predictability and can acclimatize seasonality indices as well.

The rest of the paper is structured as follows: In Section 2, country-wise confirmed and death cases datasets, preliminary of data analysis and performance evaluation are presented. The proper formulation of the proposed hybrid ARNNS and SVMAR models are discussed in Section 3. In Section 4, the experimental evaluations and results for both STM and LTM out-of-sample forecasts of global pandemic of the five countries are explained. The discussions of the results and practical implications are presented in Section 5. Finally, in Section 6, concluding remarks of this study are presented.

## 2. Data and preliminary analysis

In this study, both confirmed and death daily cases of COVID-19 for five intensely affected countries are discussed. From the Global Change Data Lab^1^ datasets are collected. The initial date of all these countries is different and the terminal date is same i.e. January 15, 2021.

### 2.1 COVID-19 datasets

All the five univariate time series datasets of both confirmed and death cases are collected for the real-time prediction purpose whereas the previous studies have forecasted only for the confirmed cases of different countries using mathematical and usual time series models. Table 1 shows the description of both confirmed and death cases with are more than 300 observations.

**Table 1:**
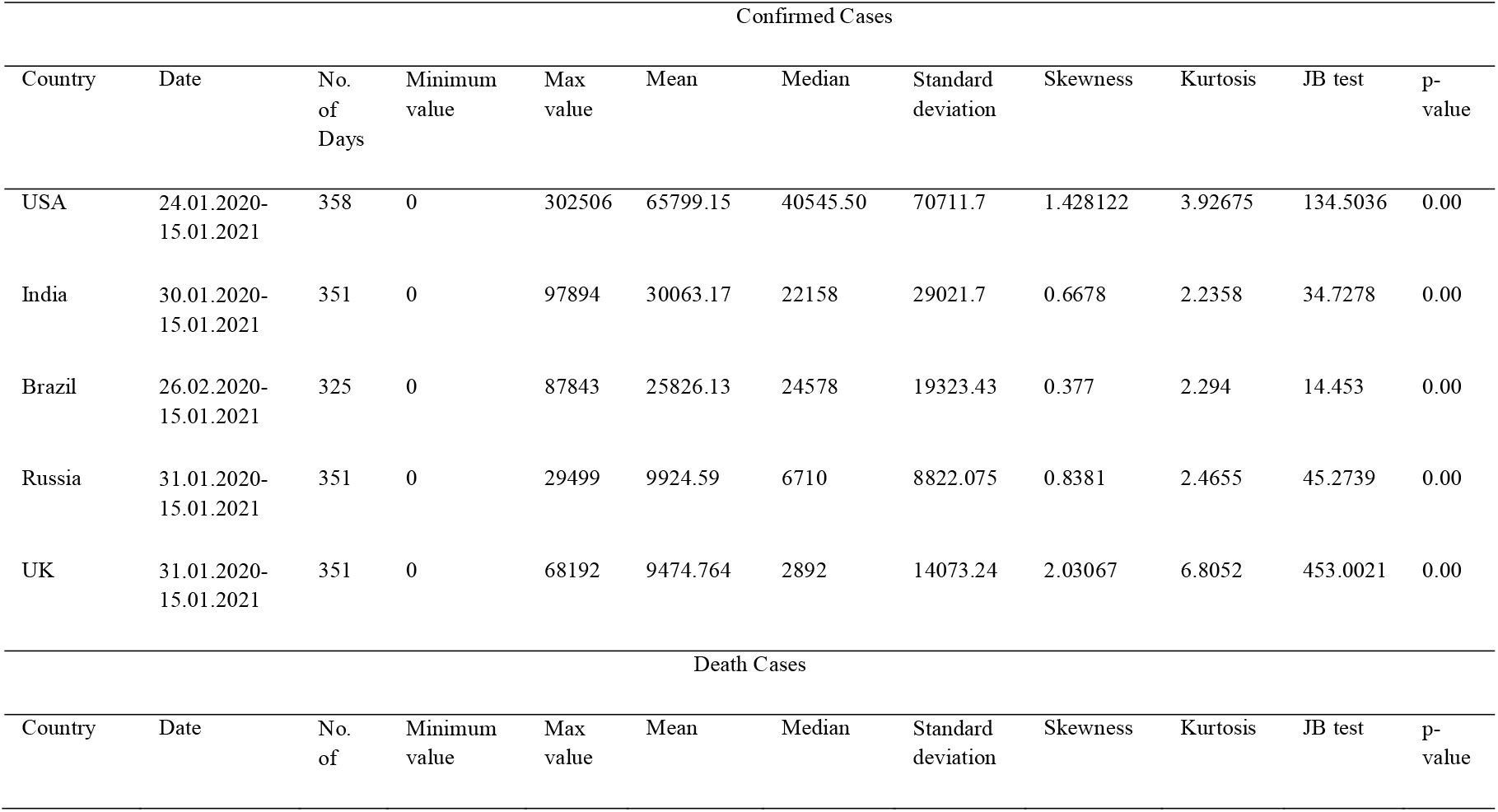
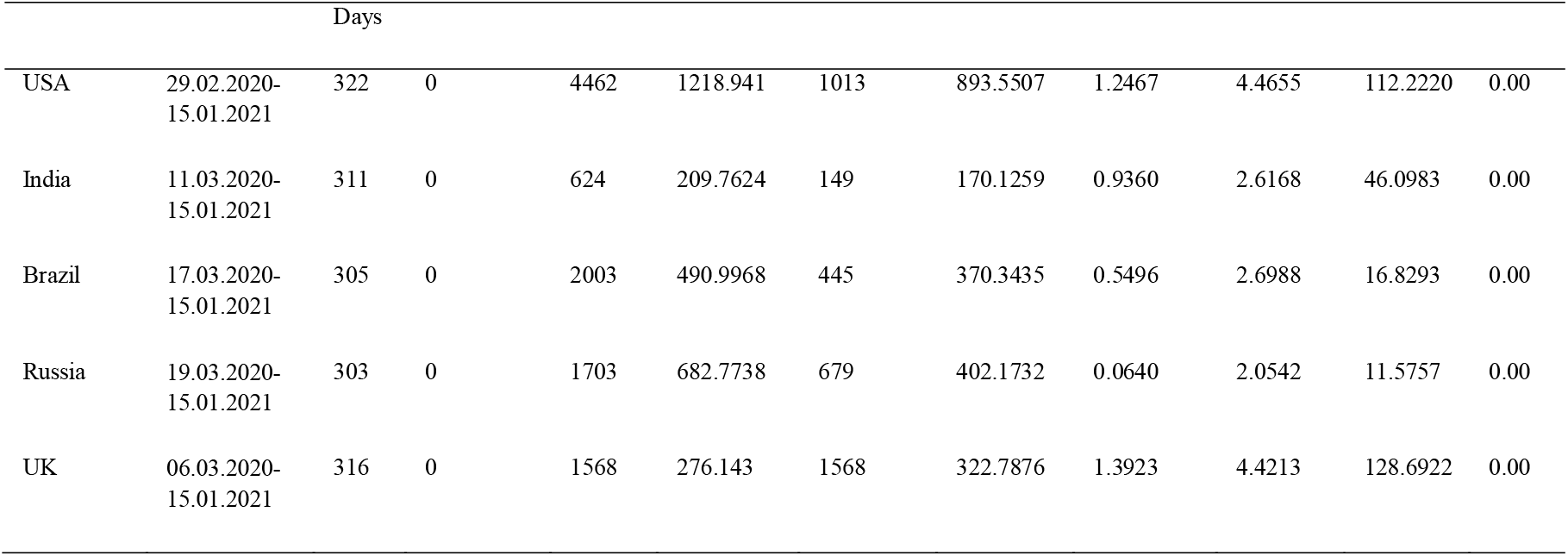
Descriptions of COVID-19 data sets of confirmed cases and death cases for USA, India, Brazil, Russia and UK respectively

### 2.2 Preliminary data analysis

Table 6.1 explains details of descriptive statistics of the COVID-19 data sets. All the COVID-19 data sets are not normally distributed. Table 4 (A-I) shows the graph of COVID-19 confirmed and death cases of five countries. These epidemic curves still not viewing the spiky dwindling nature. In this study a hybrid approach is developed and it assumes that the trend will tend to infinity in the future is considered. In the ARIMA model, the parameters *p* and *q* are determined by plotting ACF and PACF plots (see Table 4; A-I).

### 2.3 Performance Calculation

To calculate the performance of all the forecasting models under study (Ahmed et al., 2010, Chakraborty et al., 2020 and Gahan et al. 2021) MSE and MAPE are applied.

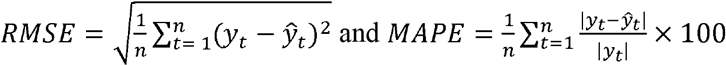

where, *y*_*t*_ : the actual value, *ŷ*_*t*_: the predicted value, and *n*: the number of data points. By definition, the lower the value of these performance values, the better is the performance of the respective forecasting model.

## 3. Methodology

Confirmed cases of COVID-19 are forecasted by applying the hybrid model of ARNN and SVM techniques (ARNNS) and of SVM and ARNN (SVMAR) techniques for predicting death cases. These proposed hybrid ARNNS and SVMAR models prevail over the deficiencies of other traditional single and hybrid models respectively. Brief descriptions of the traditional and advanced single models are given below:

### 3.1 ARNN Model

The advanced neural network ARNN (*p, k*) model is a nonlinear feed-forward neural net model with one hidden layer (having *p* lagged inputs) and *k* hidden units 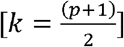 for non-seasonal data in one hidden layer (Gahan et al., 2021, Hyndman and Athanasopoulos 2018). It is designed for time series datasets which applies a pre-specified number of hidden neurons in its architecture (Faraway and Chatfield 1998). The inputs to the model are the lagged values of the datasets. Akaike Information Criterion (AIC) is used as the criterion to choose the parsimonious ARNN model by comparing different models. Using the selected past observations *y*_*t*−1_,*y*_*t*-−2_,……*y*_*t*−p_ as the inputs the estimated value *ŷ*_*t*_ is calculated. By applying a gradient descent back propagation algorithm (Rumelhart et al. 1985) the weights of the ARNN model are trained.

### 3.2 SVM Model

The Support Vector Machines (SVM) are proposed by Cortes and Vapnik (1995). It is used to have a solution of classification problems as well as non-linear regression problems. It finds the solution of the above mentioned problems by means of training the datasets. While training the sample data, it creates a hyperplane which remains within the upper and lower boundary lines. The aim of the SVM model is to find out the optimal number of hyperplanes which maximize the margin of separation between the data points. The equation of the hyperplane is *w*^*T*^ *x* + *b* = 0; where, *x*: input vector; *w*: weight which can be adjusted; *b*: bias.

### 3.3 Proposed ARNNS and SVMAR Models

Two advanced models namely ARNN and SVM models are combined to capture both non-linear and linear features of the data series (Aladag et al., 2009). By using hybrid methodology of these two models, the STM and LTM out of sample forecasting are evaluated for the univariate series. This chapter investigates the hybridization of ARNN and SVM (ARNNS) for modelling and forecasting COVID-19 confirmed cases and the hybridization of SVM and ARNN (SVMAR) for modelling and forecasting that of death cases respectively to overcome the limitations of single models and make use of their strengths. With the combination of linear and nonlinear models, the proposed methodology can assurance better performance as compared to the component models and other hybrid models.

The hybrid ARNNS model (*Z*_*t*_) can be represented as follows:

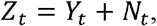

where *Y*_*t*_ is the nonlinear part and *N*_*t*_ is the linear part of the hybrid model.

Both *Y*_*t*_ and *N*_*t*_ are estimated from the data set. Let, *Y*_*t*_ be the forecast value of the ARNN model at time t and *ε*_*t*_ represent the residual at time t as obtained from the ARNN model; then *ε*_*t*_ = *Z*_*t*_ − *Ŷ*_*t*_.

The residuals are modeled by the SVM model and can be represented as follows:

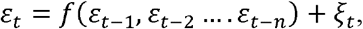

where f is a linear function and *ξ*_*t*_ is the stochastic error. The combined forecast is 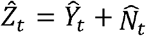 where, 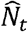 is the forecast value of the SVM model.

ARNN model captures the nonlinearity feature completely of the residual data series. So, SVM model is used which can eliminate linearity feature. There are two phases of the proposed hybrid ARNNS model. This hybrid model is able to reduce the model uncertainty, robustness, nonlinearity, non Gaussian features. The algorithm of hybrid AARNS model is explained in Table 2 (Gahan et al., 2021, Chakraborty et al., 2020).

**Table 2:**
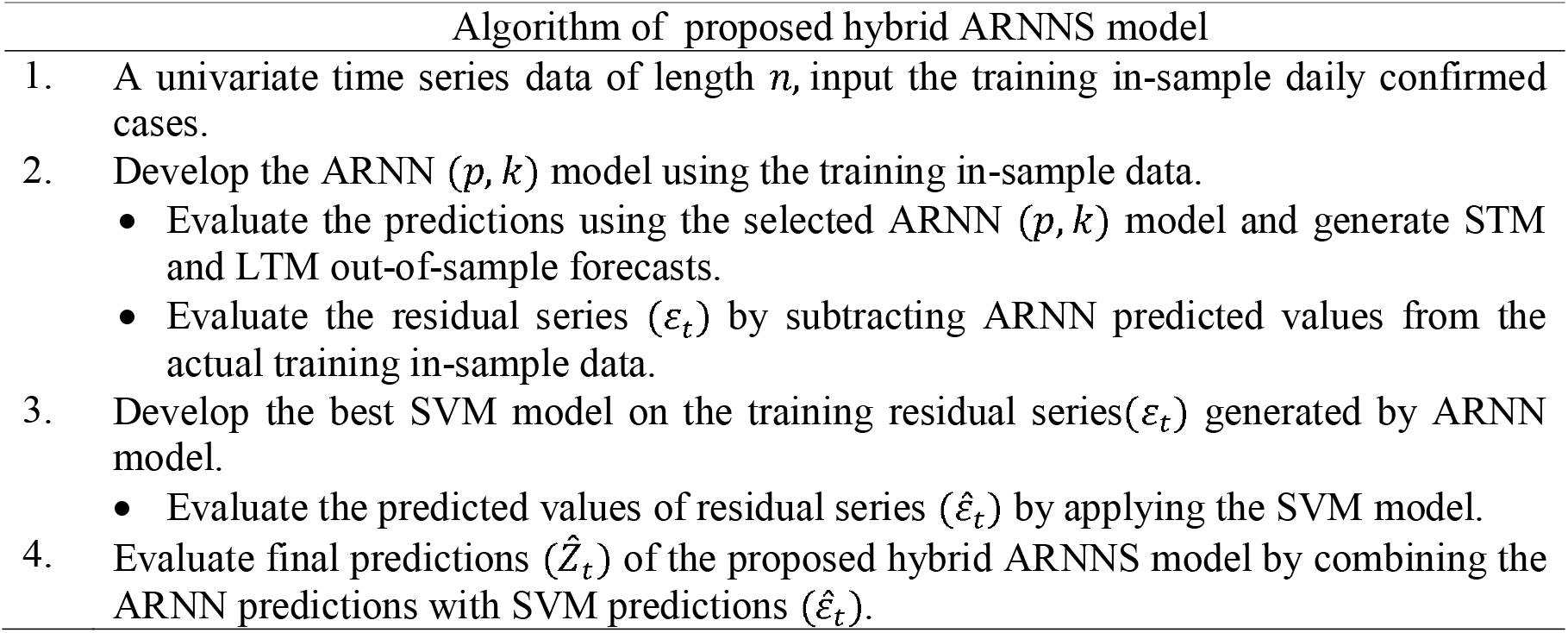
Algorithm of Proposed Hybrid ARNNS Model for Confirmed Cases of COVID-19

In the hybrid SVMAR model *Z*_*t*_ = *Y*_*t*_ + *N*_*t*,_ where *Y*_*t*_: the linear and *N*_*t*_: the nonlinear portion of this model. Evaluating *ε*_*t*_ = *Z*_*t*_ − *Ŷ*_*t*_ after estimating both these to obtain *Ŷ*_*t*_ as the forecast value of the SVM model at time t where, *ε*_*t*_ the residual value at time t. The residuals are modeled by the ARNN model and it is in the form of *ε*_*t*_ = *f*(*ε*_*t*−1_, *ε*_*t*−2_ …. *ε*_*t*−*n*_) + *ξ*_*t*_, where f is a non-linear function and *ξ*_*t*_ is the random shock. Therefore, by combing the forecast is 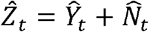, where, 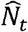 is the forecasted value of the ARNN model. As there exists autocorrelation in the residual series which SVM could not capture, for the diagnosis of the adequacy of the proposed hybrid SVMAR model is the justification to capture the residuals. So ARNN model can capture the non-linear autocorrelation relationship in the residual series. In the first phase, SVM model is applied to analyze the linear portion of the model and in the next phase, an ARNN model is employed to the residual values of the SVM model to capture the nonlinear portion. It can reduce the model uncertainty, dynamism and robustness which occur in inferential statistics and forecasting the epidemiological time series datasets. The algorithm of the proposed hybrid SVMAR model is given in Table 3 (Gahan et al. 2021 and Chakraborty et al., 2020).

**Table 3:** Algorithm of Proposed Hybrid SVMAR Model for Death Cases of COVID-19

| Algorithm of proposed hybrid SVMAR model |  |
| --- | --- |
| 1. | A univariate time series data of length $n$, input the training in-sample daily death cases. |
| 2. | Develop the SVM model using the training in-sample data.<br>• Evaluate the predictions using the selected SVM model and generate STM and LTM out-of-sample forecasts.<br>• Evaluate the residual series ($\varepsilon_t$) by subtracting SVM predicted values from the actual training in-sample data. |
| 3. | Develop the best ARNN ($p, k$) model on the training residual series ($\varepsilon_t$) generated by SVM model.<br>• Evaluate the predicted values of residual series ($\hat{\varepsilon}_t$) by applying the ARNN model. |
| 4. | Evaluate final predictions ($\hat{Z}_t$) of the proposed hybrid SVMAR model by combining the SVM predictions with ARNN predictions ($\hat{\varepsilon}_t$). |

**Table 4:**
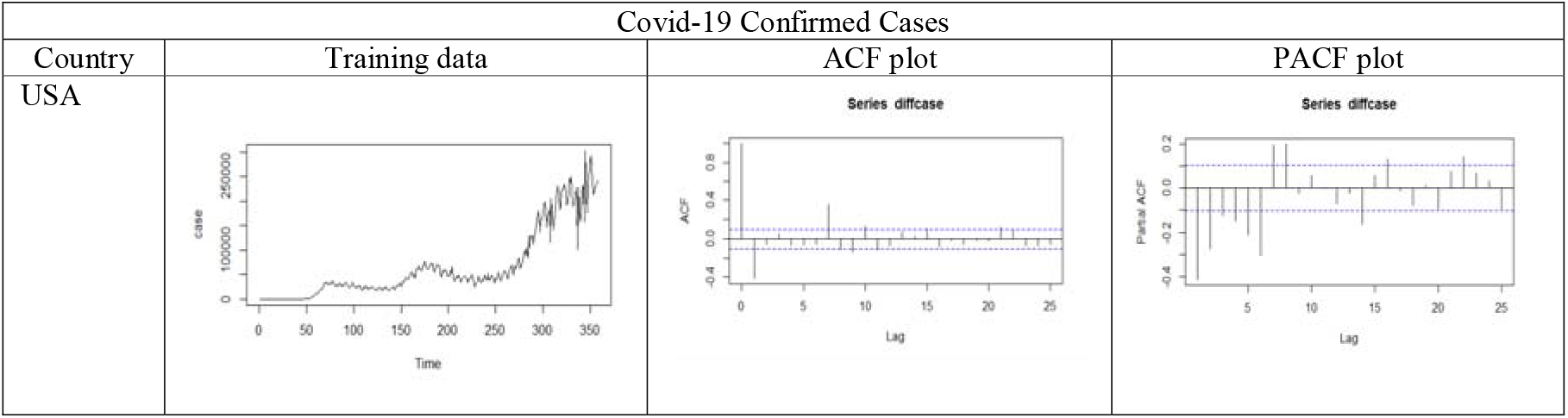
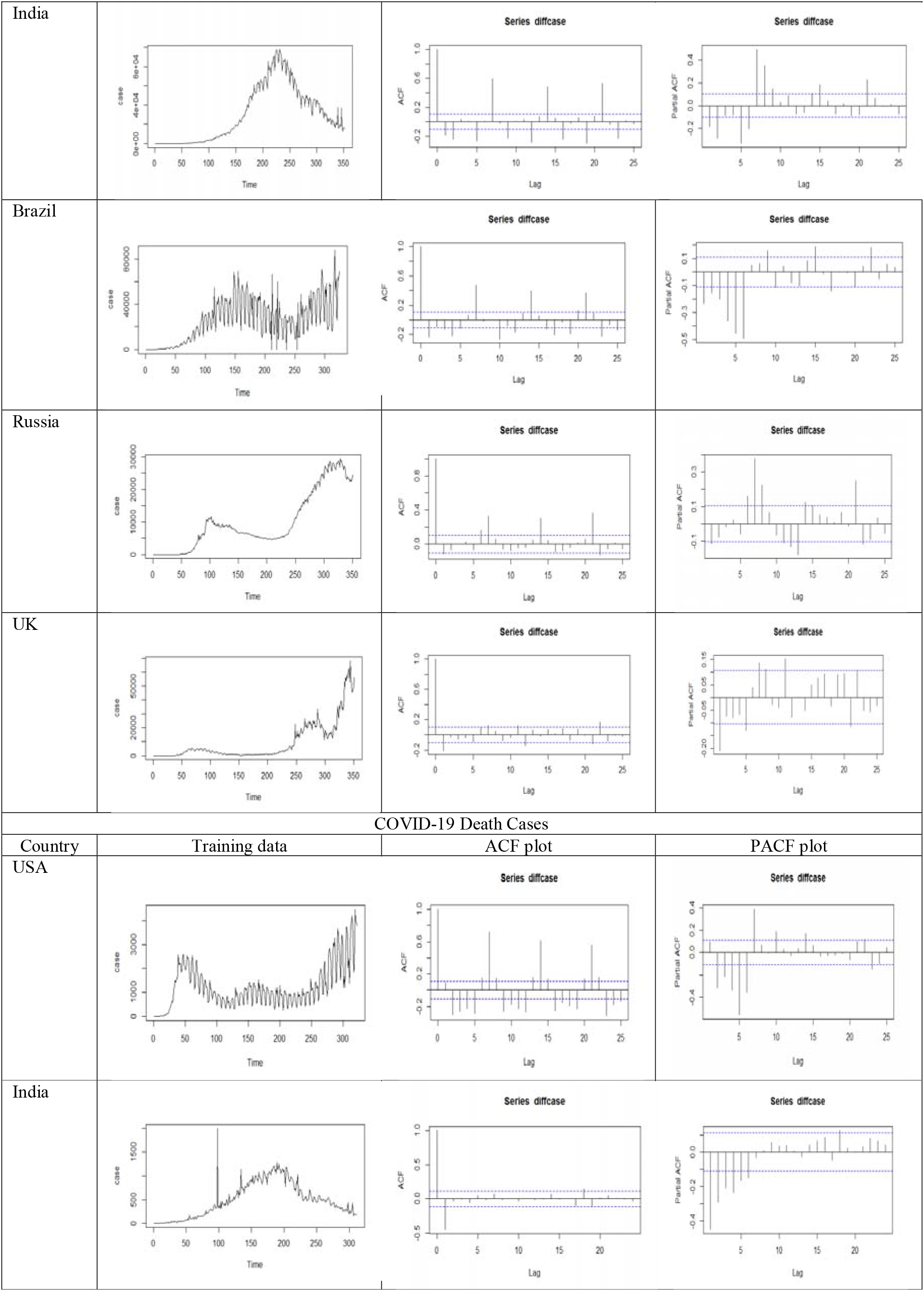
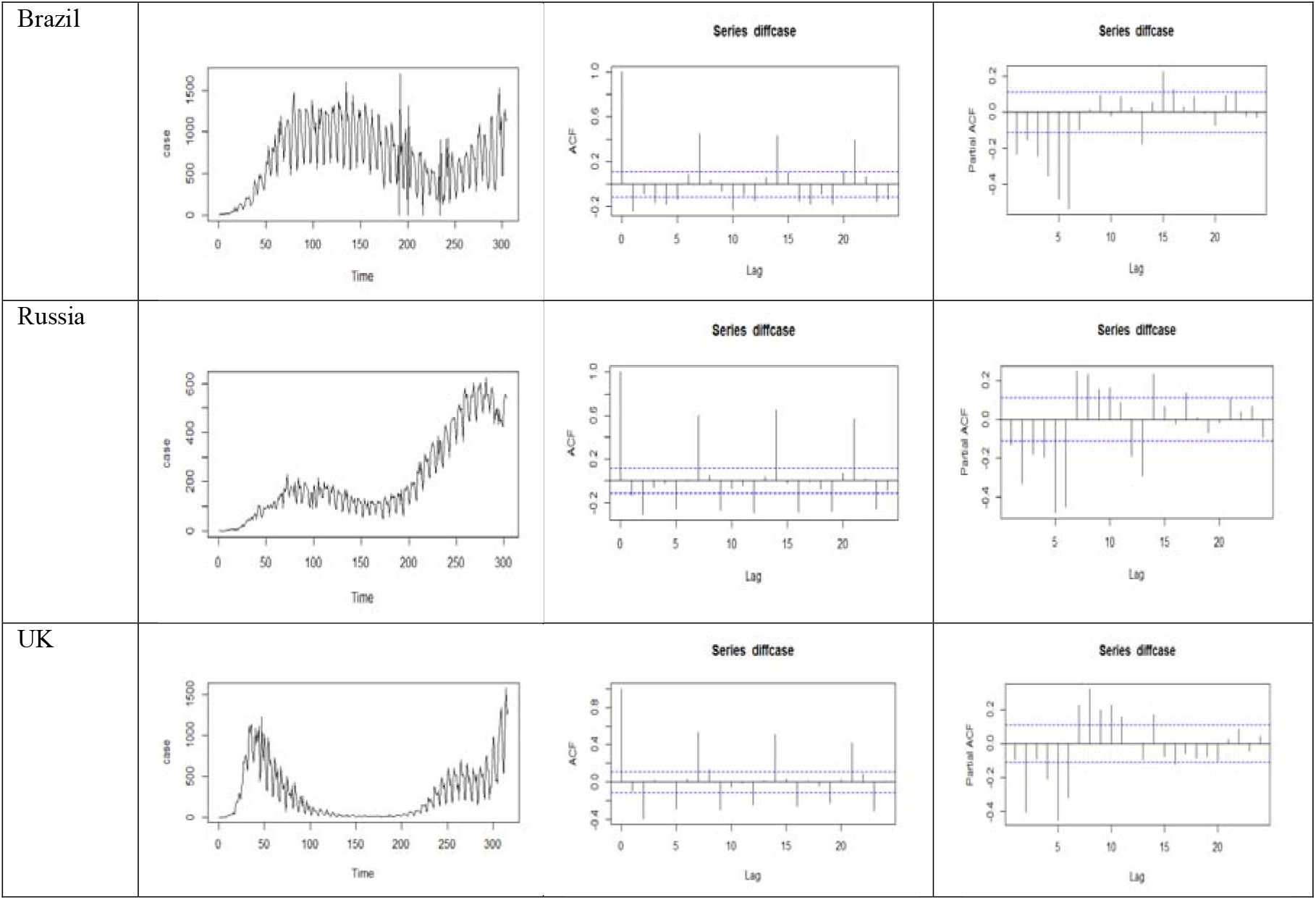
Training confirmed and death cases of COVID-19, and corresponding ACF, PACF plots for USA, India, Brazil, Russia and UK

## 4. Experimental Evaluations and Results Discussion

In presence of robust and dynamic features of the restricted COVID-19 datasets it is very difficult to predict accurately. With the accessibility of this datasets of both confirmed and death cases for five different intensely affected countries namely USA, India, Brazil, Russia and UK, prediction and forecasting of the real-time data-driven unfinished global pandemic is investigated. Two hybrid ARNNS for confirmed cases and SVMAR models for death cases are developed and applied to the above datasets respectively. These two hybrid models may be considered as the best to advise the concerned professional experts to struggle against the on-going global pandemic. For comparison purpose ARIMA, SVM, ARNN, ARIMA-ARNN, ARIMA-SVM, SVM-ARIMA and, ARNN-ARIMA models are applied to the same datasets. After analyzing statistically it is confirmed that, the datasets are nonlinear, nonstationary, and non-Gaussian in nature (see Section 2.2 and refer to Table 1). It is experimentally evaluated and compared with these two proposed hybrid ARNNS model and SVMAR (Zhang, 2003 and Chakraborty et al., 2020) through the performances of all of the above single and hybrid models (Chakraborty et al., 2020 and Gahan et al., 2021). In this chapter root mean square error (RMSE) and mean absolute percent error (MAPE) are applied to calculate the predictive performance of these models (Gahan et al., 2021, James et al., 2013 and Chakraborty et al., 2020). Since, the number of data points of COVID-19 pandemic datasets is limited thus the application of advanced deep learning techniques or other machine learning techniques will over-fit the datasets (Hastie et al., 2009 and Chakraborty et al., 2020).

The experimental evaluation of all the five datasets is applied in ARIMA, ARNN and SVM models using ‘forecast’, ‘caret’, ‘nnet’, and ‘e1071’ (Hyndman et al., 2020) applying statistical software package R. The functions like auto.arima, svm and nnetar are applied respectively in ARIMA, SVM and ARNN models. The out of sample forecasts 10-step (STM) and 100-step (LTM) ahead respectively are generated after fitting ARIMA, SVM and ARNN models. In Table 5 (A-II) plotting of the predicted and residual series of both confirmed and death cases applying ARIMA, SVM and ARNN models are shown. After fitting these three models, the residual time series, predictions of both confirmed and death cases are generated for the next ten (16^th^ Jan 2021 to 25^th^ January 2021) days and hundred (16^th^ Jan 2021 to 25^th^ April 2021) time steps ahead respectively. In Table 6 (A-III) the two proposed hybrid ARNNS and SVMAR models for actual and fittings of training data of the five countries respectively are displayed. In Table 7 and 8 (A-IV & A-V) the real-time STM and LTM out of sample forecasts (10 and 100 days ahead) for both confirmed cases and death cases using ARNN, SVM and proposed hybrid ARNNS and SVMAR models of five countries are shown respectively. For comparison purposes, all the experimental results obtained by different models are reported in Table 9 (Gahan et al., 2021 and Chakraborty et al., 2020). The performance of the proposed hybrid ARNNS and SVMAR models are superior for predicting confirmed and death cases respectively as compared to all other models. The accuracy in performance evaluation of the experimental results approves the same.

**Table 5:**
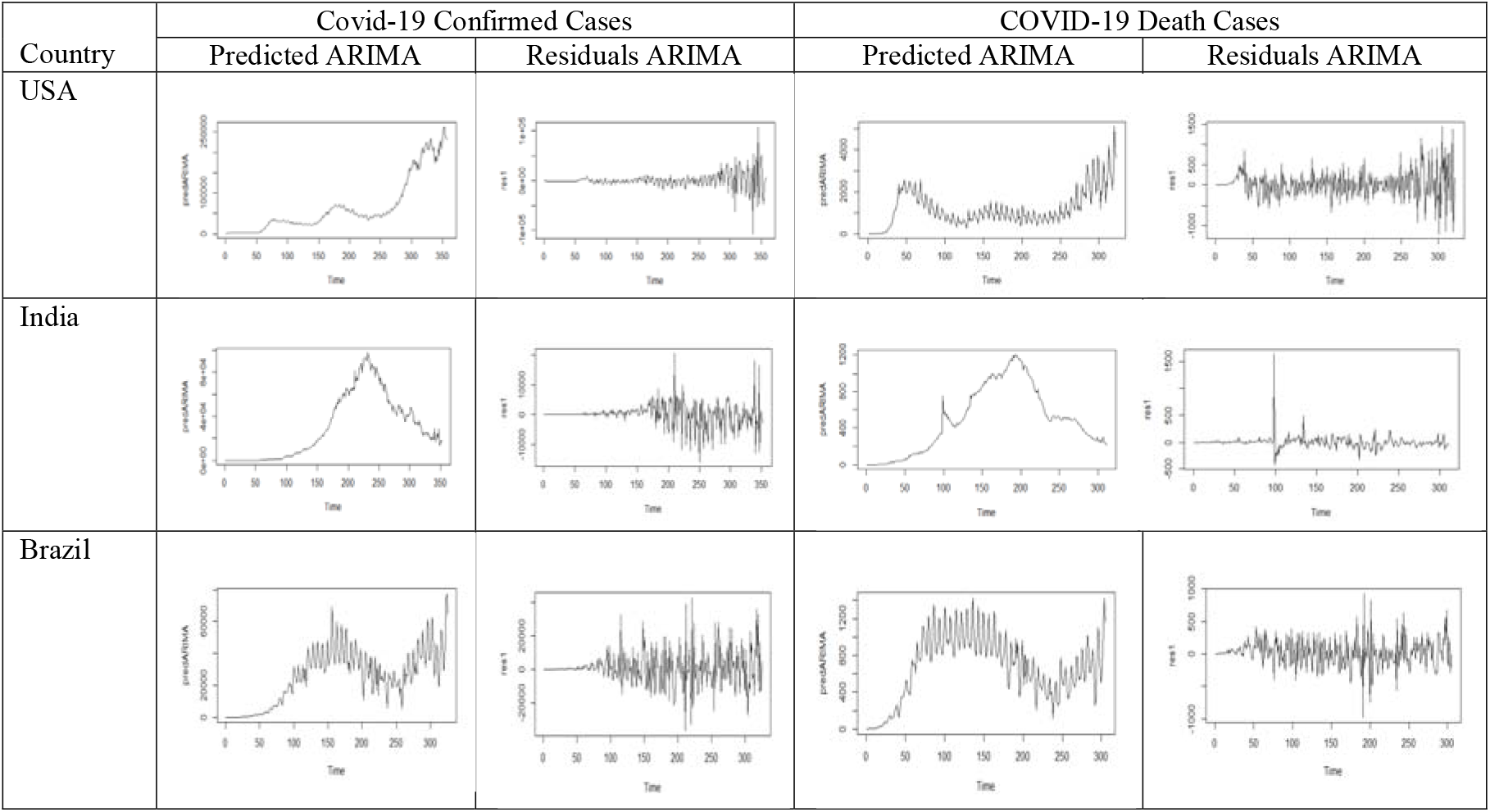
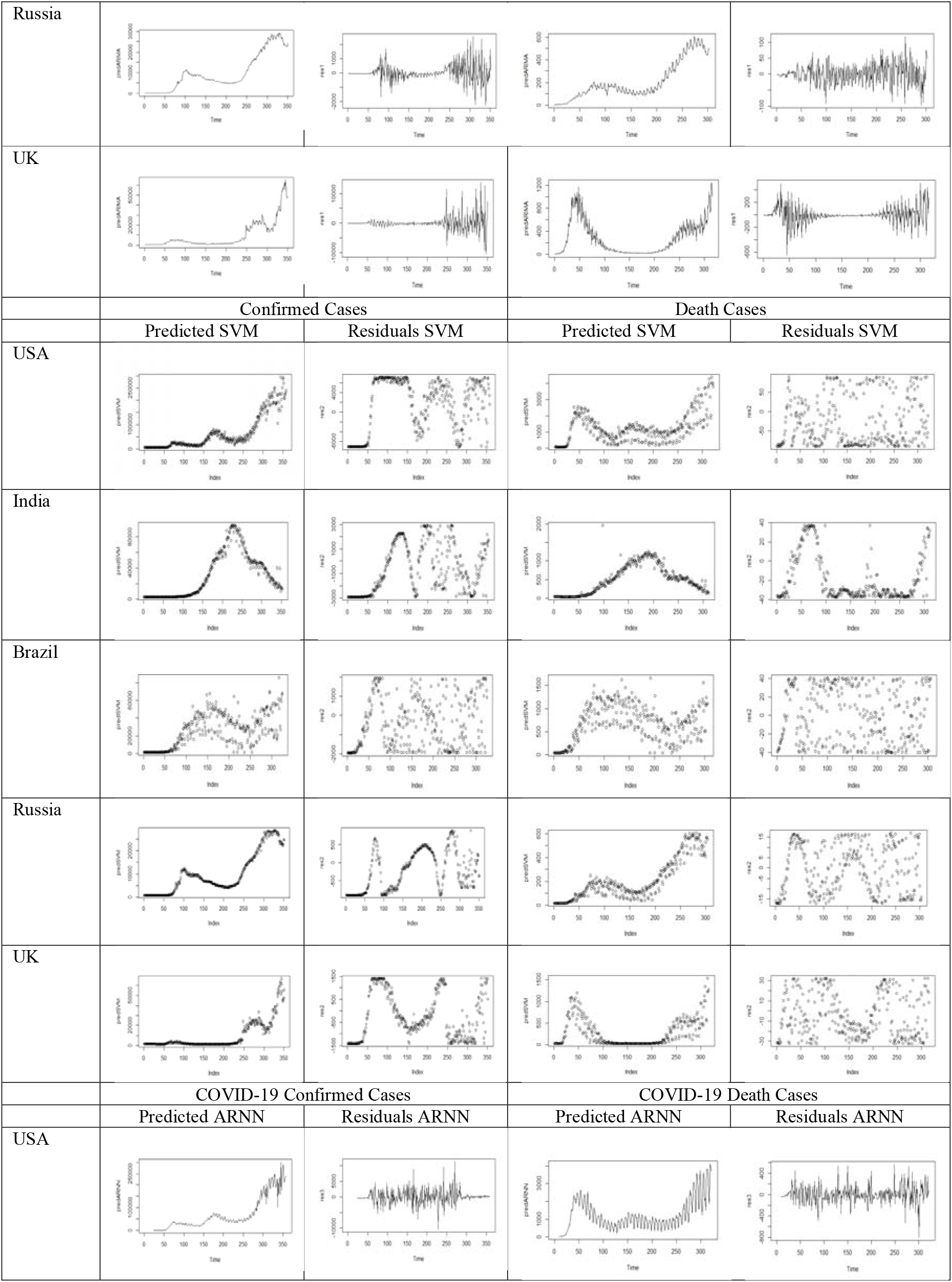
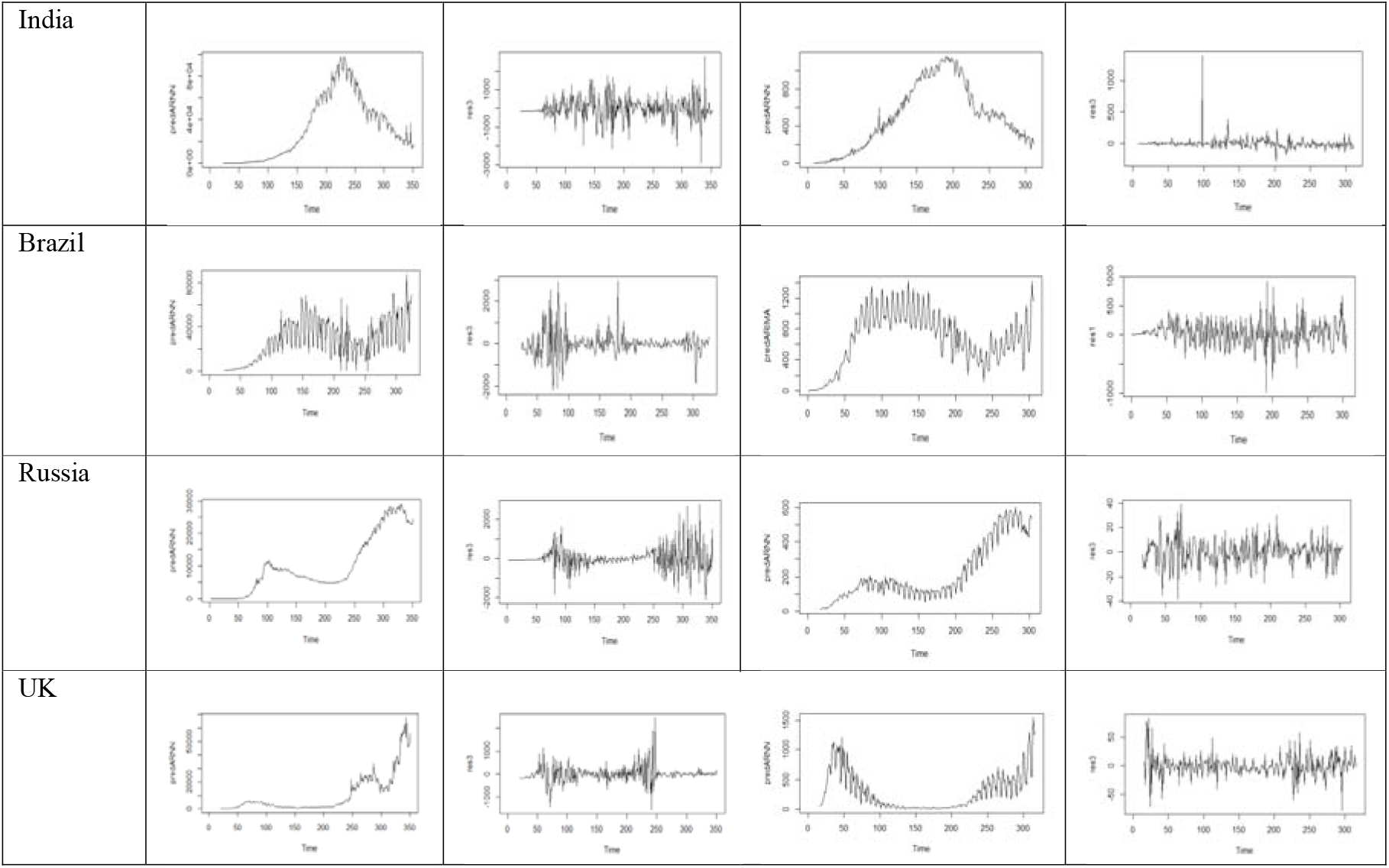
Plots of predicted and residual values of both confirmed cases and death cases of COVID-19 applying ARIMA, SVM and ARNN models for USA, India, Brazil, Russia and UK

**Table 6:**
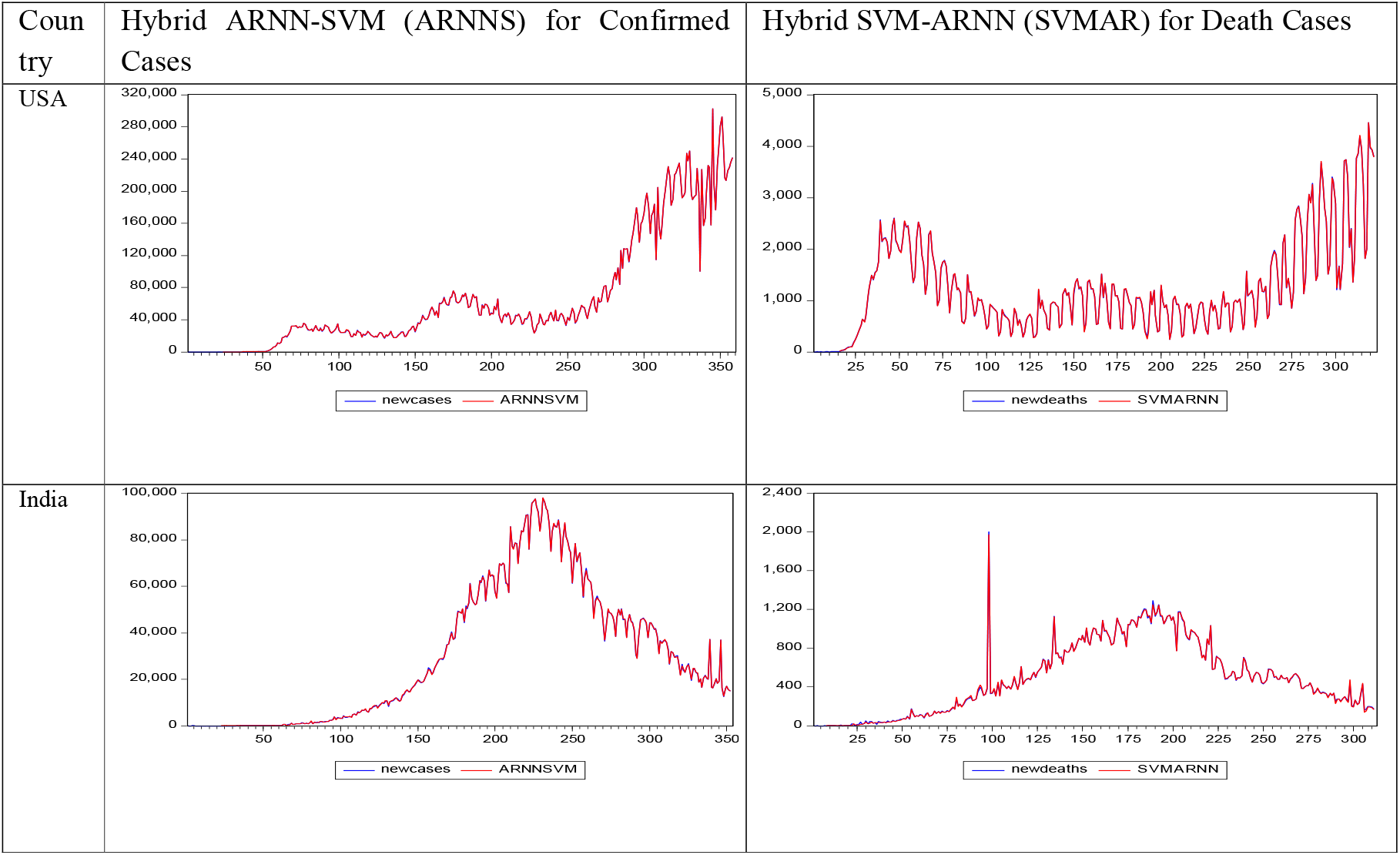
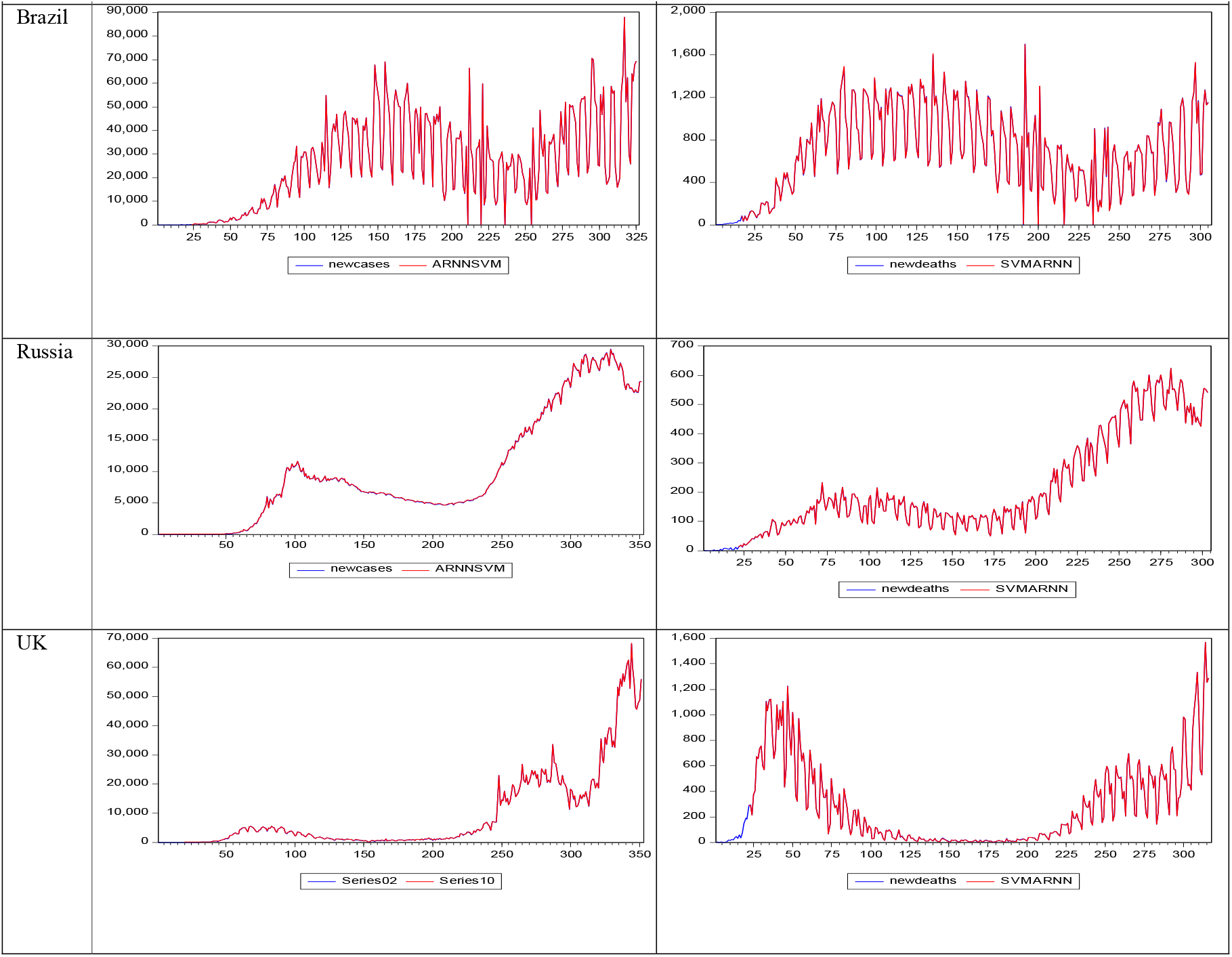
Actual and Predicted values confirmed cases applying hybrid ARNNS model and death cases applying hybrid SVMAR model for USA, India, Brazil, Russia and UK

**Table 7:**
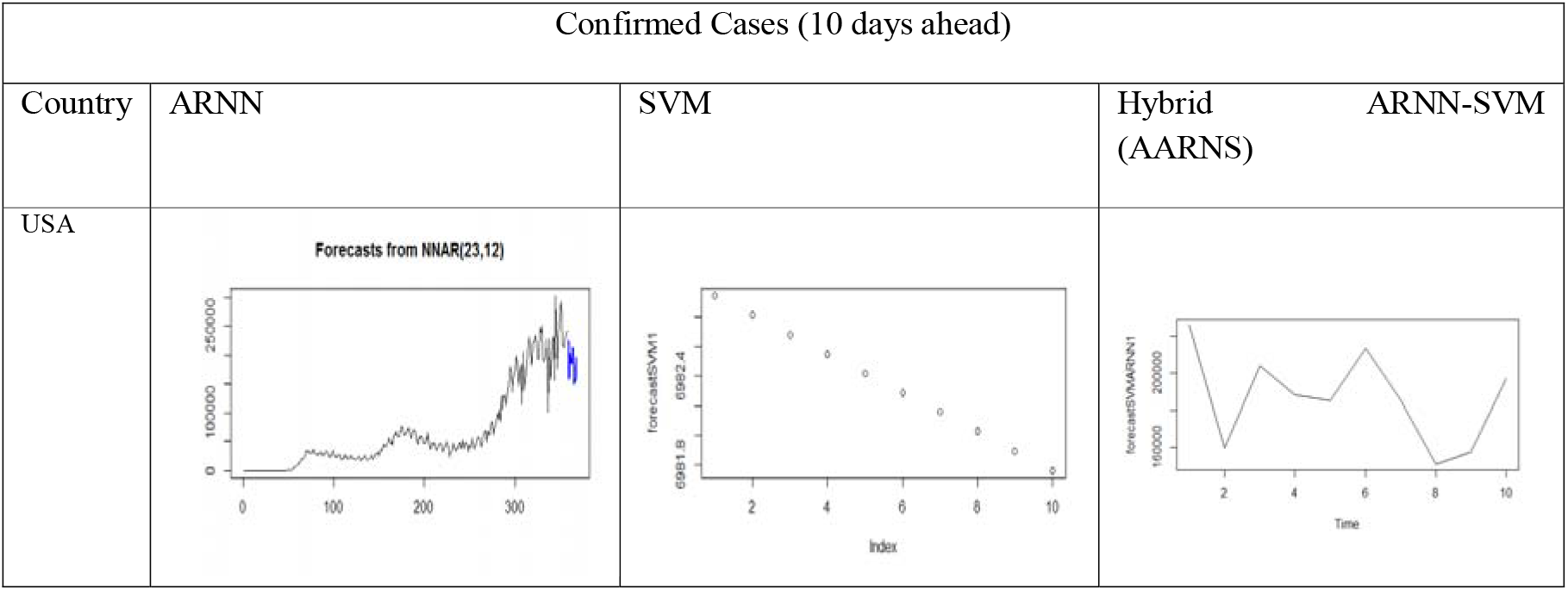
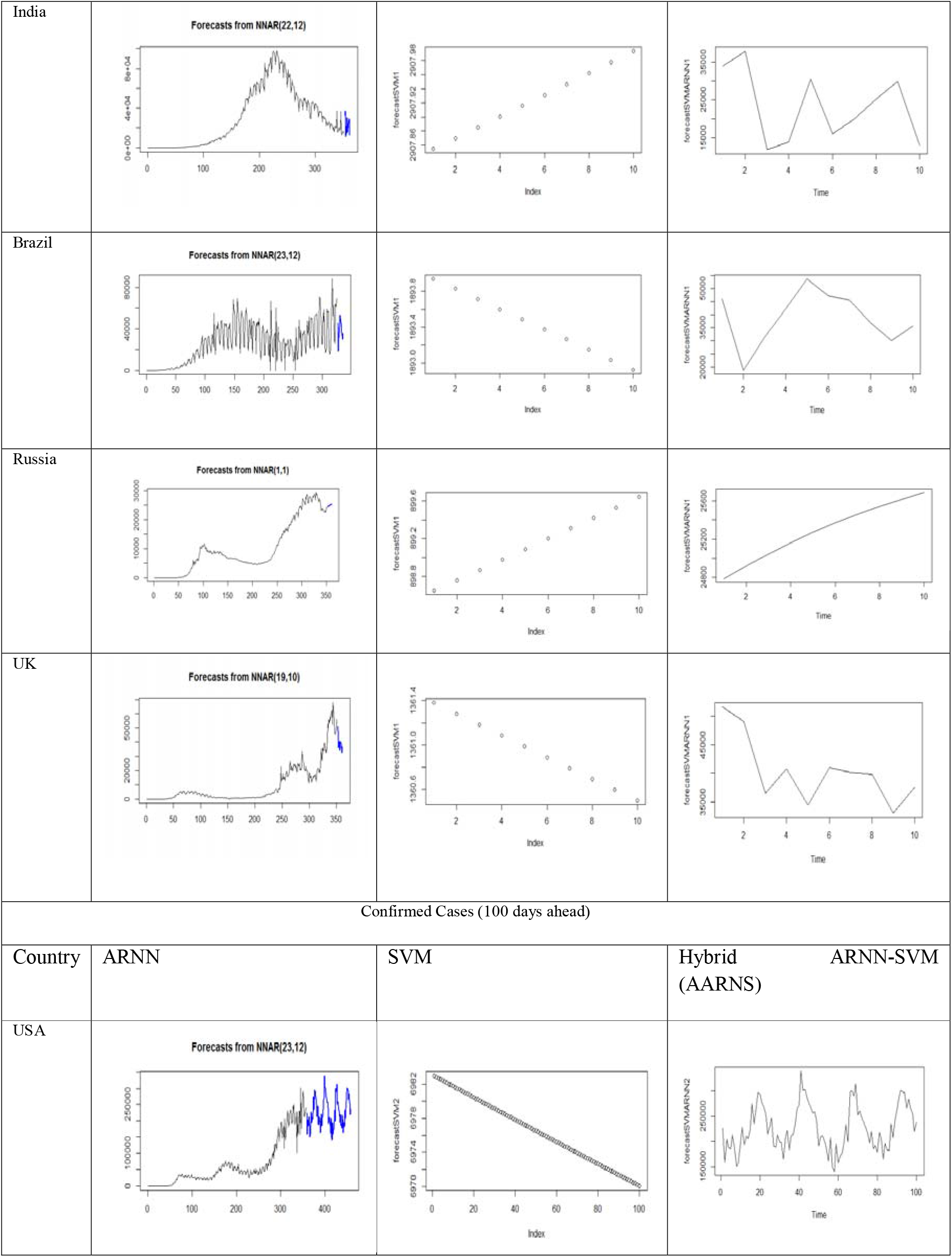
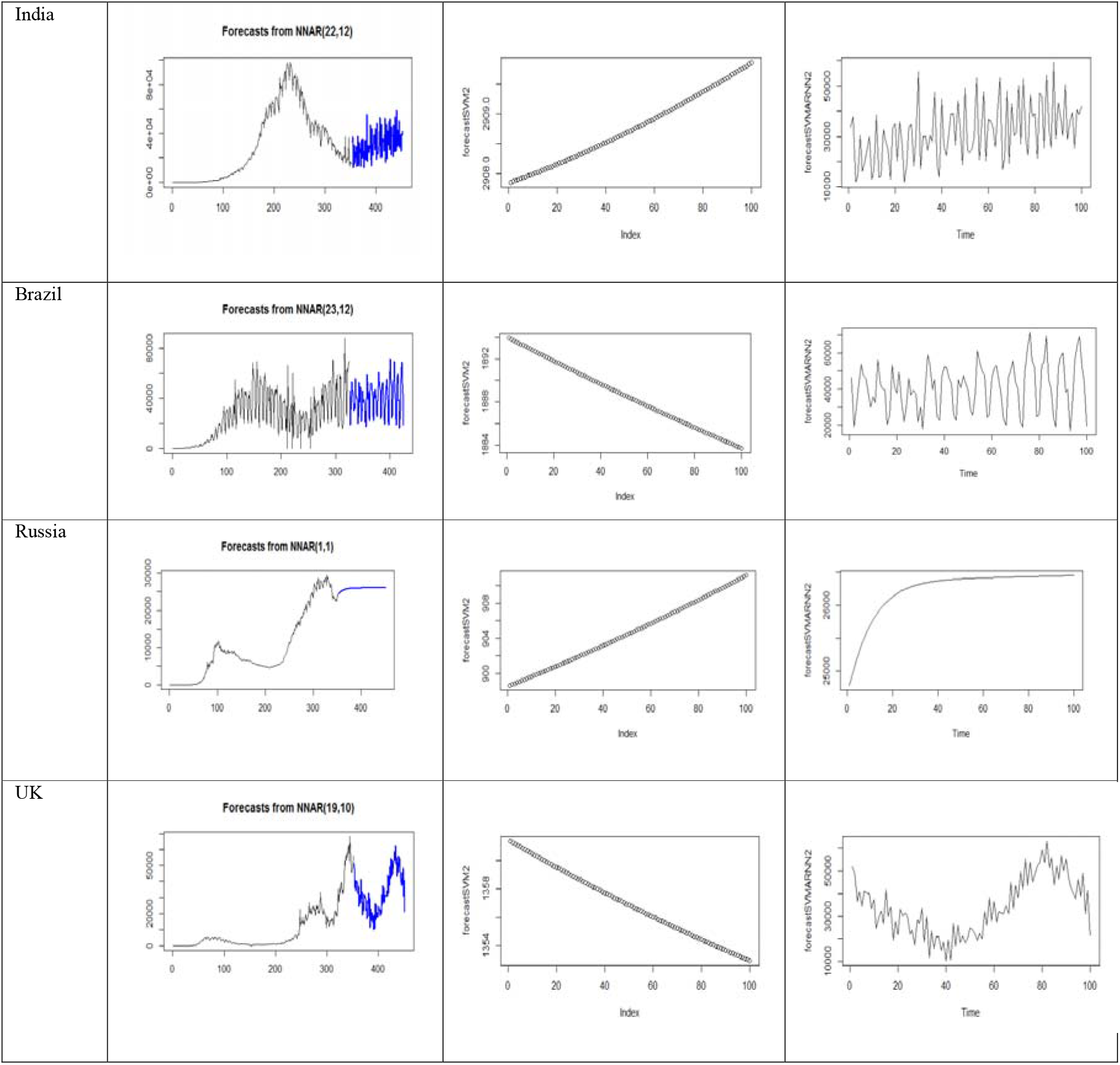
Real-time out of sample forecasts for ten and hundred days ahead of confirmed cases of COVID-19 using different forecasting models for USA, India, Brazil, Russia, and UK

**Table 8:**
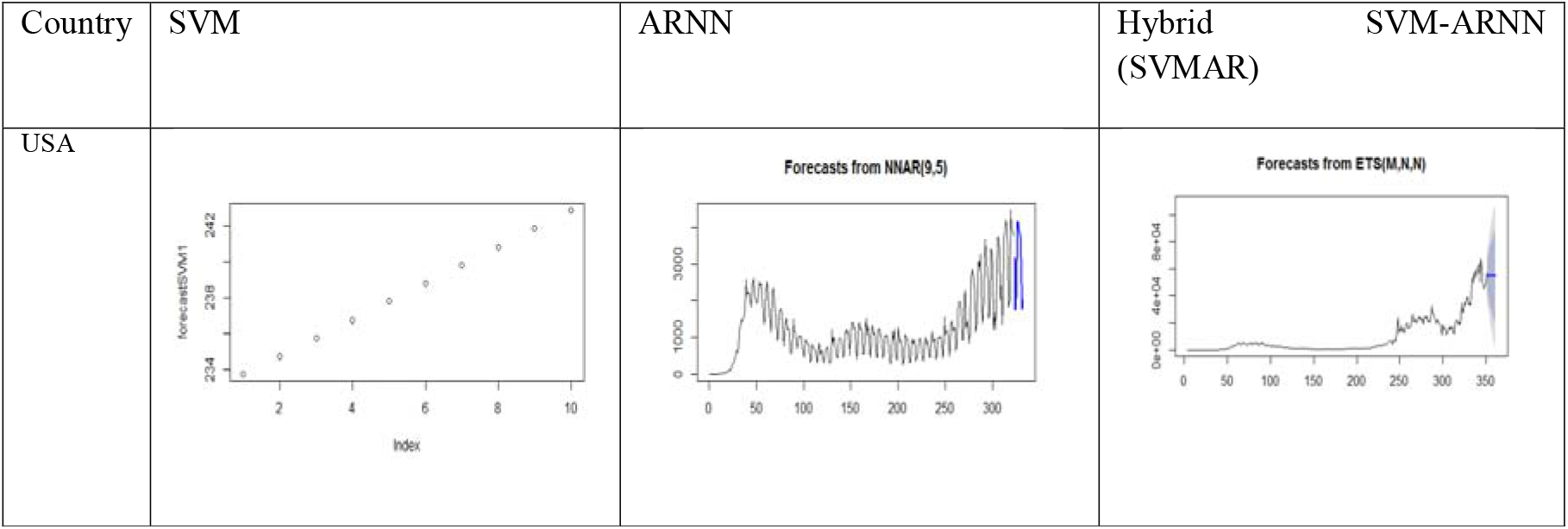
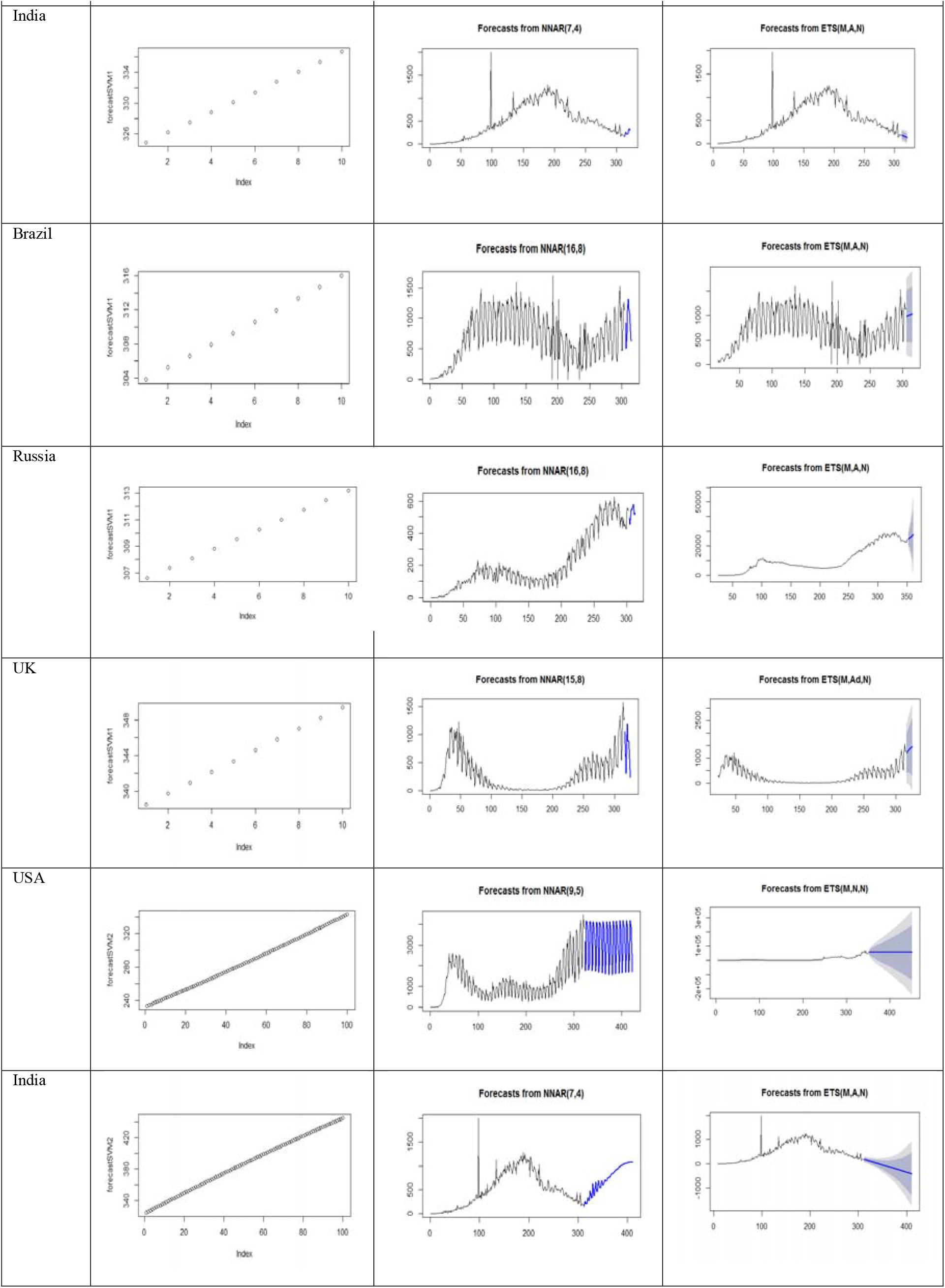
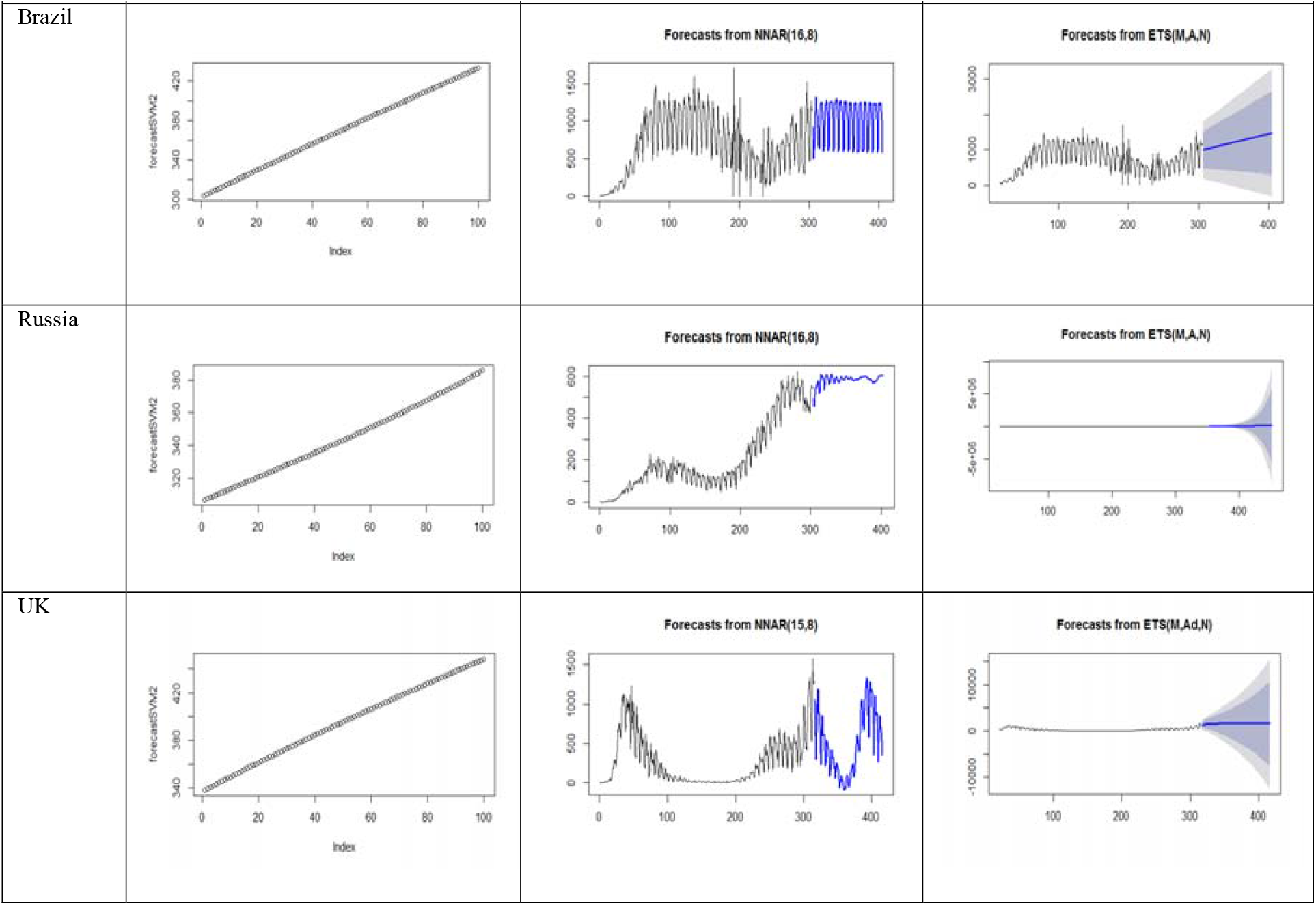
Real-time out of sample forecasts for ten and hundred days ahead death cases of COVID-19 using different forecasting models for USA, India, Brazil, Russia and UK

**Table 9:** Measures of performance for different forecasting models on five time series (training data sets only) of confirmed cases and death cases of COVID-19

| Confirmed Cases |  |  |  |  |  |  |  |  |  |  |
| --- | --- | --- | --- | --- | --- | --- | --- | --- | --- | --- |
| Country | Performance | ARIMA | SVM | ARNN | ASVM | AARNN | SVMA | SVMAR | ARNNA | ARNNS |
| USA | RMSE | 15619.6512 | 5458.9291 | 2262.0922 | 935.0941 | 1481.111 | 2159.5044 | 453.0895 | 2262.0922 | **161.2937** |
|  | MAPE | 276.1762 | 9.3437 | 59.8522 | 49.1373 | 43.9266 | **5.8543** | 8.2345 | 59.8522 | 21.8541 |
| India | RMSE | 3785.373 | 2035.561 | 636.0684 | 329.4088 | 195.8192 | 893.4736 | **166.4496** | 622.0859 | 258.1984 |
|  | MAPE | 18.7013 | 2843.066 | 158.0284 | 35.2626 | 67.2912 | 14.6277 | 29.3028 | 112.9829 | **11.7676** |
| Brazil | RMSE | 10287.2817 | 1342.1966 | 607.0248 | 781.7373 | 411.7484 | 1023.562 | 187.798 | 604.7601 | **51.1998** |
|  | MAPE | 6.9976 | 0.8501 | 0.1988 | 0.6099 | 0.1443 | 0.7844 | 0.0879 | 0.1821 | **0.0360** |
| Russia | RMSE | 588.4216 | 580.4543 | 580.6146 | 44.6095 | 39.6425 | 159.3424 | **22.7220** | 576.2854 | 49.0436 |
|  | MAPE | 20.2154 | 187.5766 | 17.1343 | 8.9053 | 4.1738 | 3.3449 | **2.4870** | 19.1337 | 11.6947 |
| UK | RMSE | 2398.178 | 1007.943 | 362.5177 | 221.9047 | 160.9041 | 346.2836 | 327.6767 | 362.5177 | **34.3085** |
|  | MAPE | 147.7804 | 855.3096 | 105.1703 | 147.4625 | 36.9326 | 47.1859 | 67.4158 | 105.1703 | **24.1865** |
| Death Cases |  |  |  |  |  |  |  |  |  |  |
| Country | Performance | ARIMA | SVM | ARNN | ASVM | AARNN | SVMA | SVMAR | ARNNA | ARNNS |
| USA | RMSE | 339.8711 | 66.1573 | 165.7598 | 25.7424 | 11.1899 | 39.4903 | **9.9614** | 166.9962 | 13.6721 |
|  | MAPE | 27.4505 | 152.183 | 35.1794 | 39.8587 | **0.5943** | 76.4975 | 1.2422 | 40.9789 | 8.4305 |
| India | RMSE | 123.1423 | 30.1295 | 101.5185 | 12.2623 | 118.262 | 10.0897 | **7.0503** | 97.7054 | 15.5105 |
|  | MAPE | 13.279 | 10.0643 | 13.3472 | 6.2619 | 14.7635 | 3.6396 | **2.7740** | 12.5621 | 5.4237 |
| Brazil | RMSE | 223.9959 | 29.2455 | 45.1709 | 17.0099 | 9.7374 | 26.2808 | **4.9802** | 48.5604 | 19.3323 |
|  | MAPE | 7.9592 | 1.2309 | 1.3996 | 0.8125 | 0.2034 | 1.1659 | **0.2023** | 1.2819 | 0.6529 |
| Russia | RMSE | 33.2443 | 11.5392 | 10.8752 | 2.5989 | 0.7812 | 8.8398 | **0.4808** | 10.9298 | 2.9233 |
|  | MAPE | 22.4764 | 22.2673 | 11.9978 | 4.3511 | 0.5342 | 9.0513 | **0.2992** | 10.8858 | 2.9096 |
| UK | RMSE | 138.8373 | 21.7596 | 19.3093 | 10.2116 | 17.8024 | 17.0800 | **1.0858** | 19.1011 | 1.4490 |
|  | MAPE | 98.4214 | 108.5191 | 36.4699 | 37.9568 | 35.8143 | 46.6345 | **4.4550** | 36.5419 | 5.5713 |

## 5. Practical Implications and Discussion

From this study it is finalised that the ARNNS and SVMAR models are forecasting well for both the cases. These two hybrid models can filter linearity by applying the ARNN and SVM models and on the whole, can explain the linearity, nonlinearity and nonstationary trends present in these datasets better as compared to other discussed models and it also yield better forecast accuracy. These results will be useful in policy and decision making for government officials to allocate sufficient health maintenance in solving to the pandemic crisis.

As it is known that all the epidemical time series data are noisy in nature because of various epidemiological factors and it is an exigent task to capture for accurate predictions. It is found that these two proposed hybrid models can predict with better precision. For all the five countries ten days STM and hundred days LTM ahead out of sample forecasts for both the cases are evaluated separately. These two hybrid models can be used as before time word of warning to fight back against this global pandemic. A list of suggestions based on the results of the real-time data-driven forecasts is:

1. Since a real-time data-driven forecast analysis is conducted, thus regular update can be made for both the cases and that of the predictions also.
2. As the out of sample forecasts follow uncertain behaviour of ten and hundred days ahead respectively. Social distancing, wearing masks, containment zone, lock down, shutdown, quarantine and sanitizing properly measures can be imposed by the government to get on the robust situations of this global pandemic.

## 6. Conclusion

It is investigated from the epidemiological data analytics, all the established and superior single like ARIMA, SVM, ARNN, and hybrid models like ASVM, AARNN, SVMA and ARNNA models are incapable to completely confine the messy behaviour of the dynamic time series datasets which contains inherent noise component. It necessitates developing both the hybrid ARNNS and SVMAR models to predict the confirmed and death cases of COVID-19 accurately and to forecast out of sample for both STM and LTM of these cases for responding to the global pandemics more powerfully.

## Data Availability

All data produced in the present study are available upon reasonable request to the authors

## Abbreviations

COVID-19: Corona Virus Disease 2019
STM: Short-term Memory
LTM: Long-term Memory
USA: United States of America
UK: United Kingdom
ARIMA: Auto Regressive Integrated Moving Average
SVM: Support Vector Machines
ARNN: Auto Regressive Neural Network
ASVM: ARIMA-SVM
AARNN: ARIMA-ARNN
SVMA: SVM-ARIMA
SVMAR: SVM-ARNN
ARNNA: ARNN-ARIMA
ARNNS: ARNN-SVM
RMSE: Root Mean Square Error
MAPE: Mean Absolute Percentage Error

## Appendix

### A-I

### A-II

### A-III

### A-IV

### A-V

## Footnotes

1 https://ourworldindia.org/coronavirus

## Notes

### Competing Interest Statement

The authors have declared no competing interest.

### Funding Statement

This study did not receive any funding

